# Pharmacogenetics of SGLT2 Inhibitors: Validation of a sex-agnostic pharmacodynamic biomarker

**DOI:** 10.1101/2023.03.07.23286875

**Authors:** Simeon I. Taylor, Hua-Ren Cherng, Zhinous Shahidzadeh Yazdi, May E. Montasser, Hilary B. Whitlatch, Braxton D. Mitchell, Alan R. Shuldiner, Elizabeth A. Streeten, Amber L. Beitelshees

**Affiliations:** Department of Medicine Division of Endocrinology, Diabetes, and Nutrition University of Maryland School of Medicine Baltimore, MD 20201 USA; Department of Radiation Oncology University of Maryland School of Medicine Baltimore, MD 20201 USA

**Keywords:** canagliflozin, diabetes, kidney, pharmacogenomics, precision medicine, sex as a biological variable, sodium-glucose cotransporter-2, type 2 diabetes, uric acid

## Abstract

**Aim:** SGLT2 inhibitors provide multiple benefits to patients with type 2 diabetes – including improved glycemic control and decreased risks of cardiorenal disease. Because drug responses vary among individuals, we initiated investigations to identify genetic variants associated with the magnitude of drug responses.

**Methods:** Canagliflozin (300 mg) was administered to 30 healthy volunteers. Several endpoints were measured to assess clinically relevant responses – including drug-induced increases in urinary excretion of glucose, sodium, and uric acid.

**Results:** This pilot study confirmed that canagliflozin (300 mg) triggered acute changes in mean levels of several biomarkers: fasting plasma glucose (−4.1 mg/dL; p=6x10), serum creatinine (+0.05 mg/dL; p=8×10^-4^), and serum uric acid (−0.90 mg/dL; p=5×10^-10^). The effects of sex on glucosuria depended upon how data were normalized. Whereas males’ responses were ∼60% greater when data were normalized to body surface area, males and females exhibited similar responses when glucosuria was expressed as grams of urinary glucose per gram-creatinine. The magnitude of glucosuria was not significantly correlated with fasting plasma glucose, estimated GFR, or age in these healthy non-diabetic individuals with estimated GFR>60 mL/min/1.73m^2^.

**Conclusions:** Normalizing data relative to creatinine excretion will facilitate including data from males and females in a single analysis. Furthermore, because our ongoing pharmacogenomic study (NCT02891954) is conducted in healthy individuals, this will facilitate detection of genetic associations with limited confounding by other factors such as age and renal function.

**Registration:** NCT02462421 (clinicaltrials.gov)

**Funding:** Research grants from the National Institute of Diabetes and Digestive and Kidney Diseases: R21DK105401, R01DK108942, T32DK098107, and P30DK072488.

## 1 INTRODUCTION

Although head-to-head comparative effectiveness studies demonstrate that diabetes drugs are not strongly differentiated with respect to mean HbA1c-lowering ^1^, there is wide variation in individual responses to the same therapy. A recent head-to-head study reported HbA1c- lowering efficacies (mean ± SEM) of 0.89% ± 0.24% for canagliflozin and 1.44% ± 0.39% for liraglutide ^2^. This corresponds to standard deviations of ∼0.9% and ∼1.5%, respectively. While some patients experienced little if any decrease in HbA1c, others experienced >2.5% HbA1c- lowering. This inter-individual variation results from a combination of genetic and environmental factors. Pharmacogenetics has potential to provide insights enabling physicians to prescribe therapies for individual patients based on predictors of individual responses and risks of adverse effects ^1^.

Pharmacogenomics has identified genetic variants contributing to inter-individual variation in responses to diabetes drugs ^3–7^. Some genetic variants alter pharmacokinetics by altering function of drug transporters or drug metabolizing enzymes ^3, 8^. Other genetic variants alter functions of proteins that mediate drug responses, thereby altering pharmacodynamics ^4, 6, 9^. This pilot study represents one step toward identifying genetic variants contributing to inter-individual variation in responses to SGLT2 inhibitors – an increasingly important class of diabetes drugs ^10, 11^. Glucuronidation is the principal pathway whereby SGLT2 inhibitors are metabolized. Loss-of-function variants in glucuronidation enzymes alter pharmacokinetics of SGLT2 inhibitors by increasing drug exposure ^8^, but these variants are unlikely to cause major alterations in drug responses in routine clinical use because SGLT2 inhibitors are usually administered at maximally effective doses. Increasing drug exposure may not alter responses to drugs that are administered at maximally effective doses.

Several approaches have been applied in pharmacogenomic research – including acute studies in healthy volunteers assessing pharmacodynamic endpoints ^3, 12^ or chronic studies in disease patients assessing clinical endpoints such as HbA1c or cardiovascular outcomes ^9, 12^. Short-term studies in healthy volunteers offer methodological advantages – including minimization of confounding factors due to co-existing diseases and effects of changing co-medications during the study. This pilot study provided an opportunity to validate pharmacodynamic endpoints for our ongoing GWAS of pharmacodynamic responses to canagliflozin (NCT02891954). Our approach was inspired by high priority NIH initiatives: (a) scientific rigor and reproducibility ^13^; (b) investigation of sex as a biological variable ^14^; and (c) precision medicine ^15^.

Research participants collected two separate 24-hour urine collections. We confirmed the reproducibility of urine collections as indicated by absence of a statistically significant difference in creatinine content between the two separate urine collections – a critical prerequisite for our twin objectives of minimizing the impact of random variation and maximizing the impact of genetic variation. This pilot study also investigated the impact of participants’ self-designated sex on pharmacodynamic responses. Mean canagliflozin-induced glucosuria was substantially greater in males than females when expressed as grams of glucose per 1.73 m^2^ of body surface area or per kg of body weight. In contrast, mean magnitudes of canagliflozin-induced glucosuria were similar in males and females when expressed as grams of glucose per gram of creatinine. Accordingly, our ongoing pharmacogenomic study will normalize drug-induced glucosuria data relative to urinary creatinine excretion so that the magnitude of canagliflozin-induced glucosuria would be relatively independent of sex (“sex-agnostic”).

## 2 METHODS

### 2.1 Study population

The Old Order Amish population of Lancaster County, PA immigrated to the Colonies from Central Europe in the early 1700’s. There are currently ∼40,000 Old Order Amish individuals in Lancaster County, PA – nearly all of whom trace their ancestry back ∼15 generations to ∼750 founders ^16–18^. Investigators at the University of Maryland Baltimore have investigated genetic determinants of cardiometabolic health in this population since 1993. These studies generated a genotype database used to compile a list of individuals to be invited to participate in this clinical trial. Individuals with any of four homozygous genotypes were eligible to participate: (a) nonsense mutation in *SLC5A4* (rs62239058); (b) nonsense mutation in *SLC5A9* (rs850763); (c) missense variant in *SLC2A*9 (rs1689079); and (d) “wild type” major alleles of *SLC5A4*, *SLC5A9*, and *SLC2A9*.

### 2.2 Conduct of clinical trial (NCT02462421)

This clinical trial was reviewed and approved by the University of Maryland Baltimore’s Institutional Review Board. A research nurse accompanied by a member of the Amish community made home visits to invite selected individuals to participate in the study. Potential participants were invited to provide informed consent after nurses explained the study in detail. Additional methodological details are summarized in the Supplementary Appendix.

### 2.3 Statistical analyses

Student’s t-tests as implemented in Microsoft Excel and GraphPad Prism software were used to assess treatment effects (paired t-test) and differences between groups (unpaired t-tests). Nominal p-values are presented without correcting for multiple comparisons. A nominal p-value of <0.05 was taken as the threshold for statistical significance.

## 3 RESULTS

### 3.1 Disposition and baseline characteristics of participants

This clinical trial assessed pharmacodynamic responses of thirty healthy Old Order Amish participants to a single dose of canagliflozin (300 mg). Disposition of the participants is summarized in Fig. S1. The study population (17 females/13 males) had a mean age of 57.8 ± 2.5 years and a mean BMI of 28.0 ± 0.9 kg/m^2^ (Table S1). Baseline laboratory data are summarized in Table S1.

### 3.2 Responses to canagliflozin: ι fasting plasma glucose, ι serum uric acid, and η serum creatinine

Canagliflozin inhibits SGLT2, thereby decreasing proximal tubular reabsorption of glucose and sodium ^10^. Drug-induced glucosuria triggered a ∼4.5% decrease in mean fasting plasma glucose levels (p=0.00006; Fig. 1A). The decrease in proximal tubular sodium reabsorption induced natriuresis (Table 1, Fig. S2), which triggered a modest volume contraction as reflected by an ∼8% increase in serum creatinine (p=0.0008; Fig. 1B). Canagliflozin also increased urinary excretion of uric acid, leading to a ∼21% decrease in mean serum uric acid levels (p=5 x 10^-10^; Fig. 1C) ^10, 19–21^. Thus, administration of a single dose of canagliflozin exerted the expected pharmacodynamic effects on circulating biomarkers in healthy volunteers – thereby serving as positive controls for our clinical trials (NCT02462421 and NCT02891954).

**Figure 1.**
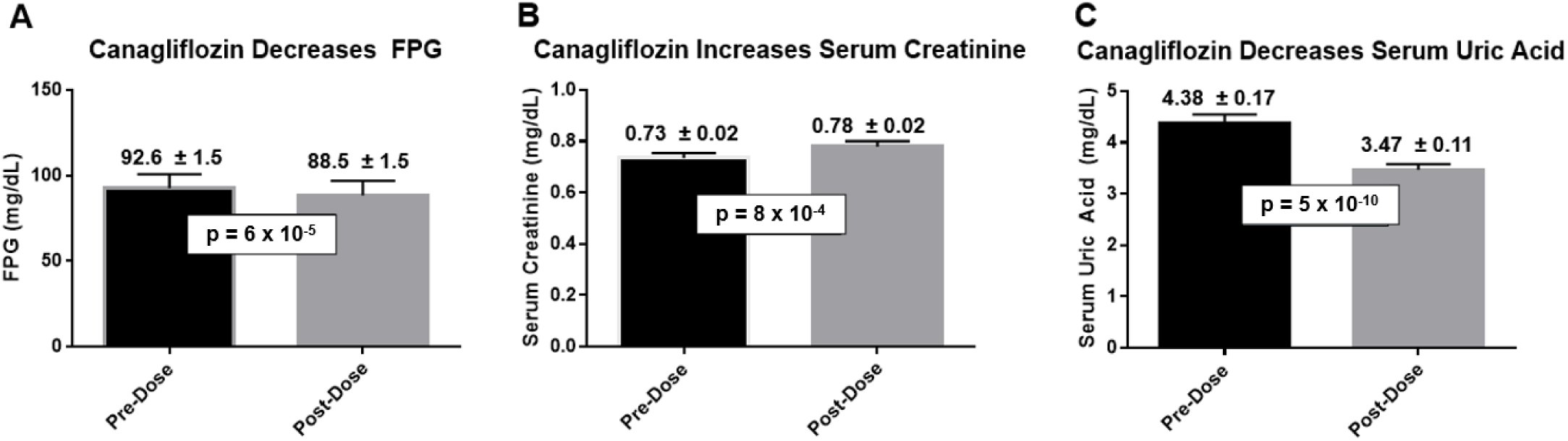
Acute effects of canagliflozin on circulating biomarkers. Baseline blood samples were obtained for measurement of fasting plasma glucose (panel A), serum creatinine (panel B), and serum uric acid (panel C). Canagliflozin (300 mg, p.o.) was administered 24 hours later to each of the participants. Twenty-four hours after administration of canagliflozin, fasting blood samples were obtained to assess the impact of canagliflozin on fasting plasma glucose, serum creatinine, and serum uric acid. Data are presented as mean ± SEM (N=30).

**Table 1.**
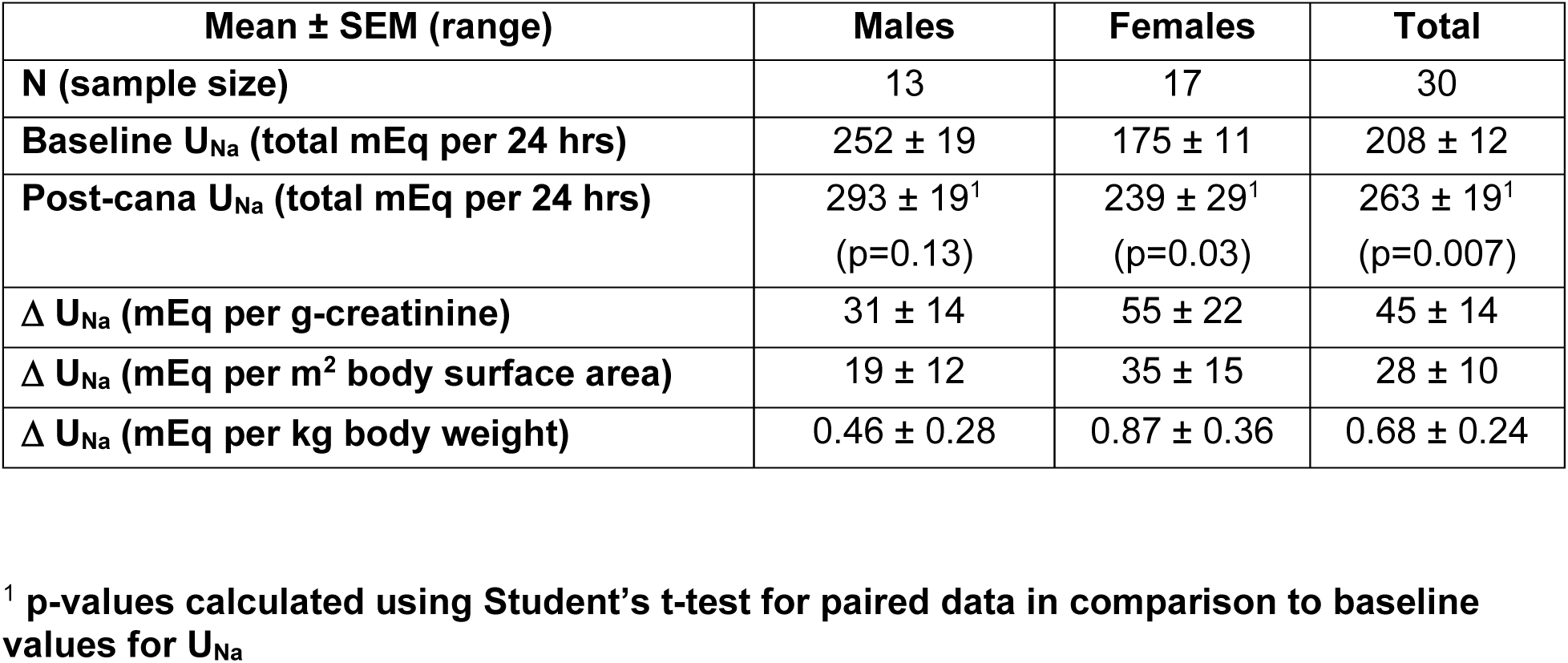
Canagliflozin increases urinary sodium excretion (U_Na_).

| **Mean ± SEM (range)** | **Males** | **Females** | **Total** |
| --- | --- | --- | --- |
| **N (sample size)** | 13 | 17 | 30 |
| **Baseline U<sub>Na</sub> (total mEq per 24 hrs)** | 252 ± 19 | 175 ± 11 | 208 ± 12 |
| **Post-cana U<sub>Na</sub> (total mEq per 24 hrs)** | 293 ± 19<sup>1</sup><br>(p=0.13) | 239 ± 29<sup>1</sup><br>(p=0.03) | 263 ± 19<sup>1</sup><br>(p=0.007) |
| **Δ U<sub>Na</sub> (mEq per g-creatinine)** | 31 ± 14 | 55 ± 22 | 45 ± 14 |
| **Δ U<sub>Na</sub> (mEq per m<sup>2</sup> body surface area)** | 19 ± 12 | 35 ± 15 | 28 ± 10 |
| **Δ U<sub>Na</sub> (mEq per kg body weight)** | 0.46 ± 0.28 | 0.87 ± 0.36 | 0.68 ± 0.24 |

<sup>1</sup> p-values calculated using Student's t-test for paired data in comparison to baseline values for U<sub>Na</sub>

Our primary efficacy endpoint is based on measurements of biomarkers in 24-hour urine collections. We, therefore, investigated the reproducibility of 24-hour urine collections. On average, participants excreted 1286 ± 81 mg of creatinine during the 24 hours before canagliflozin as compared to 1252 ± 74 mg of creatinine during the 24 hours after canagliflozin administration (Fig. S3A). The quantities of 24-hour urinary creatinine before and after canagliflozin administration were highly correlated with one another (r=0.95; p=10^-15^). When we compared creatinine contents of the two specimens collected by each individual participant, differences were <5% for ∼75% of paired urine collections (Fig. S3B). These data demonstrate that most participants successfully obtained complete collections of urine produced during a 24-hour time period.

### 3.3 A sex-agnostic approach to normalizing urinary biomarker data

Urinary excretion of glucose provides a quantitative biomarker for efficacy of canagliflozin ^22–27^. Furthermore, enhanced urinary glucose excretion plays a critical role in mediating two important benefits provided by SGLT2 inhibitors: improved glycemic control in diabetic patients and weight loss in overweight/obese patients ^10^. To enable pharmacogenomic investigation, it is critical to optimize the approach to normalize data in order to compare glucosuric responses among participants. We considered two standard approaches: (a) normalizing urinary glucose excretion relative to body size (i.e., body weight or body surface area); or (b) normalizing glucosuria relative to 24-hour urinary creatinine excretion. Body weight was tightly correlated with body surface area (r=0.99) in both males and females (Fig. S4A), suggesting that these two parameters provide comparable indices for body size. Although 24-hour urinary creatinine was correlated with body surface area (r=0.68 for males and r=0.65 for females), males on average excreted ∼60% more creatinine than females (Fig. S4B) when compared at the same body surface area. Creatinine is produced by a non-enzymatic chemical reaction – i.e., the conversion of phosphocreatine to creatinine ^28^. Because phosphocreatine is located primarily in lean tissues (e.g., muscle), the rate of creatinine production implicitly reflects lean tissue mass. When glucosuria is expressed relative to urinary excretion of creatinine, this implicitly indexes glucosuria relative to lean body mass.

Glucosuria was correlated with urinary creatinine excretion in both males (r=0.65; Fig. 2A) and females (r=0.86; Figs. 2C). Glucosuria was also correlated with body surface area (Figs. 2B and 2D) or body weight (Fig. S5) – albeit with smaller correlation coefficients (0.29-0.33 in males and 0.54-0.55 in females). The closer correlations with urinary creatinine excretion favors the approach of normalizing glucosuria relative to urinary creatinine excretion. Furthermore, this approach yielded similar mean values for glucosuria in both sexes: 35.4 ± 2.0 in males versus 39.4 ± 1.4 g-glucose per g-creatinine in females (p=0.12; Fig. 3B). In contrast, mean values for glucosuria were 37-39% larger in males than females when expressed as a function of body surface area (Fig. 3A) or body weight (Fig. 3C). Thus, because mean values of glucosuria are similar in both sexes when expressed on a per g-creatinine basis, this will greatly facilitate including data from both sexes in the same analysis.

**Figure 2.**
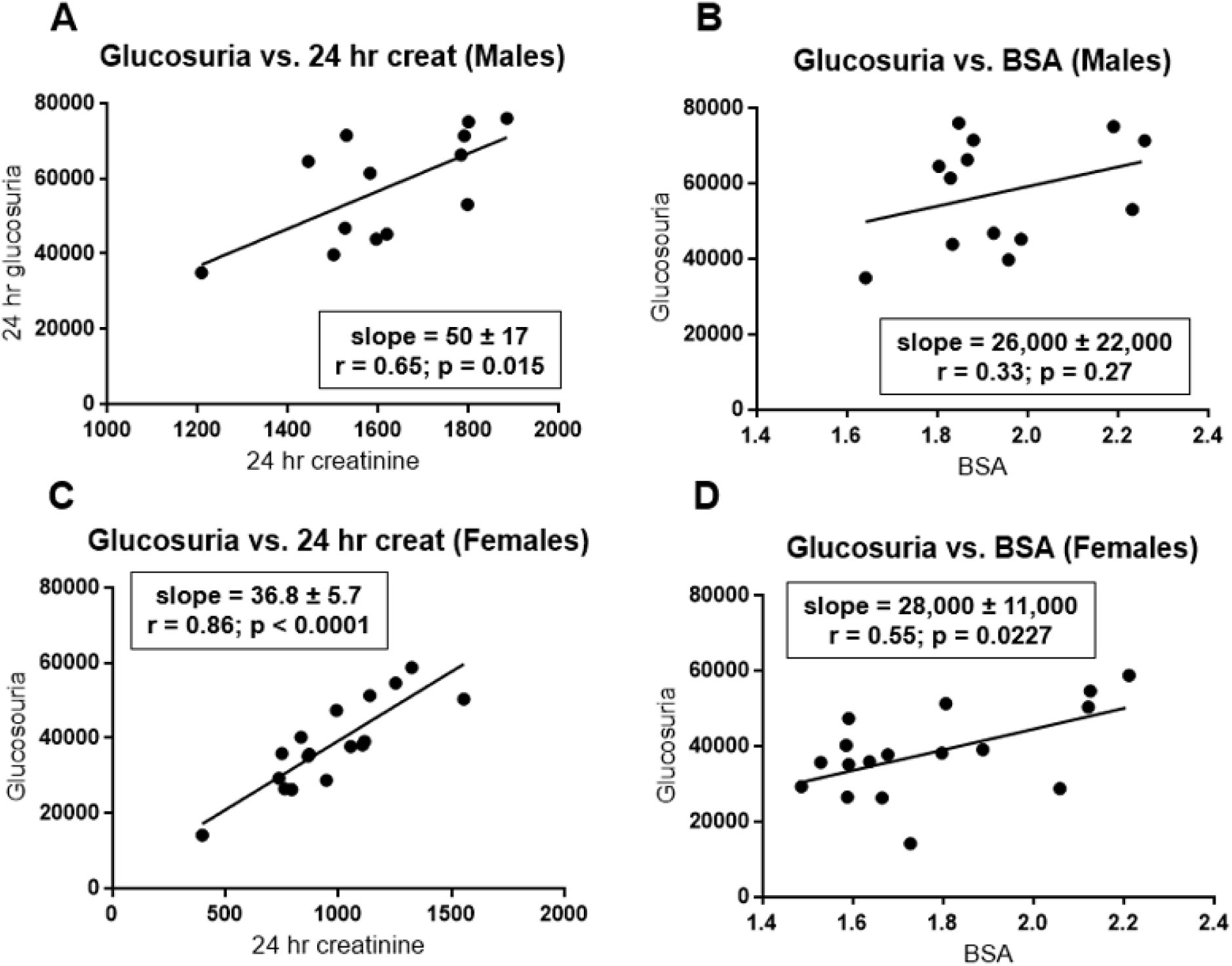
Correlations of glucosuria with urinary creatinine excretion and body surface area. Glucosuria (expressed as mg/day) is plotted as a function of either 24-hour urinary creatinine excretion (mg/day) (panels A and C) or body surface area (m^2^) (panels B and D). Data are presented separately for males (panels A and B) and females (panels C and D). Using data analysis programs provided in Excel, we estimated slopes for the least-square lines and correlation coefficients. P-values were calculated using GraphPad Prism software.

**Figure 3.**
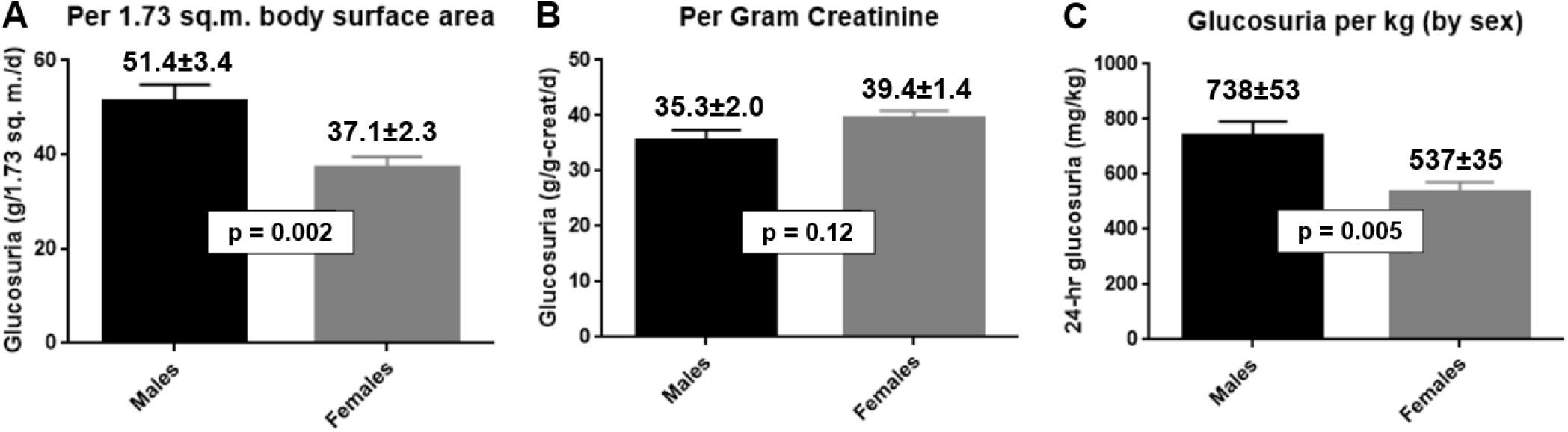
Sex differences with respect to the magnitude of canagliflozin-induced glucosuria. Using data presented in Figure 3, we calculated mean ± SEM for canagliflozin-induced 24-hour urinary glucose excretion normalized in one of two ways: grams of glucose per 1.73 m^2^ body surface area (Panel A) or grams of glucose per gram of creatinine (Panel B). Data for male and female participants are represented as either black or gray columns, respectively.

### 3.4 Inter-individual variation in canagliflozin-induced glucosuria

We observed substantial inter-individual variation in the magnitude of canagliflozin-induced increases in urinary glucose excretion. The magnitude of glucosuria varied over an approximately twofold range: from ∼25 to ∼50 g-glucose/g-creatinine (Fig. S6). Two critical factors contribute to determining the quantity of glucose filtered at the glomerulus (the “filtered glucose load”): (a) plasma glucose levels and (b) glomerular filtration rates. Thus, we investigated correlations of glucosuria with fasting plasma glucose levels (FPG), creatinine clearance rates, and age (Fig. S7). This clinical trial focused on healthy volunteers – excluding patients with HbA1c ≥ 6.5% or eGFR < 60 mL/min/1.73 m^2^. In this population, we did not observe statistically significant correlations of 24-hour urinary excretion of glucose with fasting plasma glucose, measured creatinine clearance rates, or age (Fig. S7). In addition to being statistically insignificant, the correlation coefficients were quite small; variances in fasting plasma glucose, creatinine clearance, and age accounted for only ∼1.4%, 0.5%, and 4.8% of the total variance in canagliflozin-induced glucosuria, respectively. Measured creatinine clearance was closely correlated with eGFR in both males and females (Fig. S8). However, we focused on the measured creatinine clearance because it is likely to be a more valid index of each individual’s glomerular filtration rate.

### 3.5 Canagliflozin-induced natriuresis

Because SGLT2 functions as a co-transporter for glucose plus Na^+^, canagliflozin inhibits proximal tubular reabsorption of both glucose and Na^+ 10^. Drug-induced natriuresis was calculated by subtracting 24-hour urinary Na^+^ excretion observed at baseline from 24-hour urinary Na^+^ excretion observed in the 24 hours after administration of canagliflozin. Canagliflozin increased mean urinary Na^+^ excretion by ∼25% in the total population (N=30; p=0.007) (Table 1). Research participants were free to eat *ad libitum* and to engage in their usual daily activities. As no effort was made to control sodium intake, there may have been substantial day-to-day variation in the sodium content of participants’ diets and/or the quantity of sodium lost through sweating. Such day-to-day variation represent probable sources of unmeasured confounders with respect to estimates of drug-induced natriuresis. Notwithstanding these caveats, estimates of drug-induced natriuresis appeared to be larger for females than males – regardless of whether the magnitude of 24-hour Na^+^ urinary excretion was normalized relative to urinary creatinine excretion, body surface area, or body weight (Table 2).

### 3.6 Canagliflozin-induced uricosuria

SGLT2 inhibitors enhance urinary excretion of uric acid and decrease serum levels of uric acid ^10, 19–21^. Consistent with these previous publications, we observed that canagliflozin induced a 2- to 10-fold increase in fractional excretion of uric acid. The magnitude of the increase in fractional excretion of uric acid was correlated with the magnitude of the decrease in serum uric acid levels (r=0.48; p=0.006) (Fig. 4A). Although mechanisms mediating SGLT2 inhibitor-induced uricosuria have not been established, the effect may be an indirect consequence of the increase in glucose concentrations in the renal tubular fluid ^19^. Accordingly, we investigated whether the increase in fractional excretion of uric acid was correlated with canagliflozin-induced glucosuria. However, we did not detect statistically significant correlation between the drug-induced increase in fractional excretion of uric acid and canagliflozin-induced glucosuria or natriuresis (Fig. 4BC).

**Figure 4.**
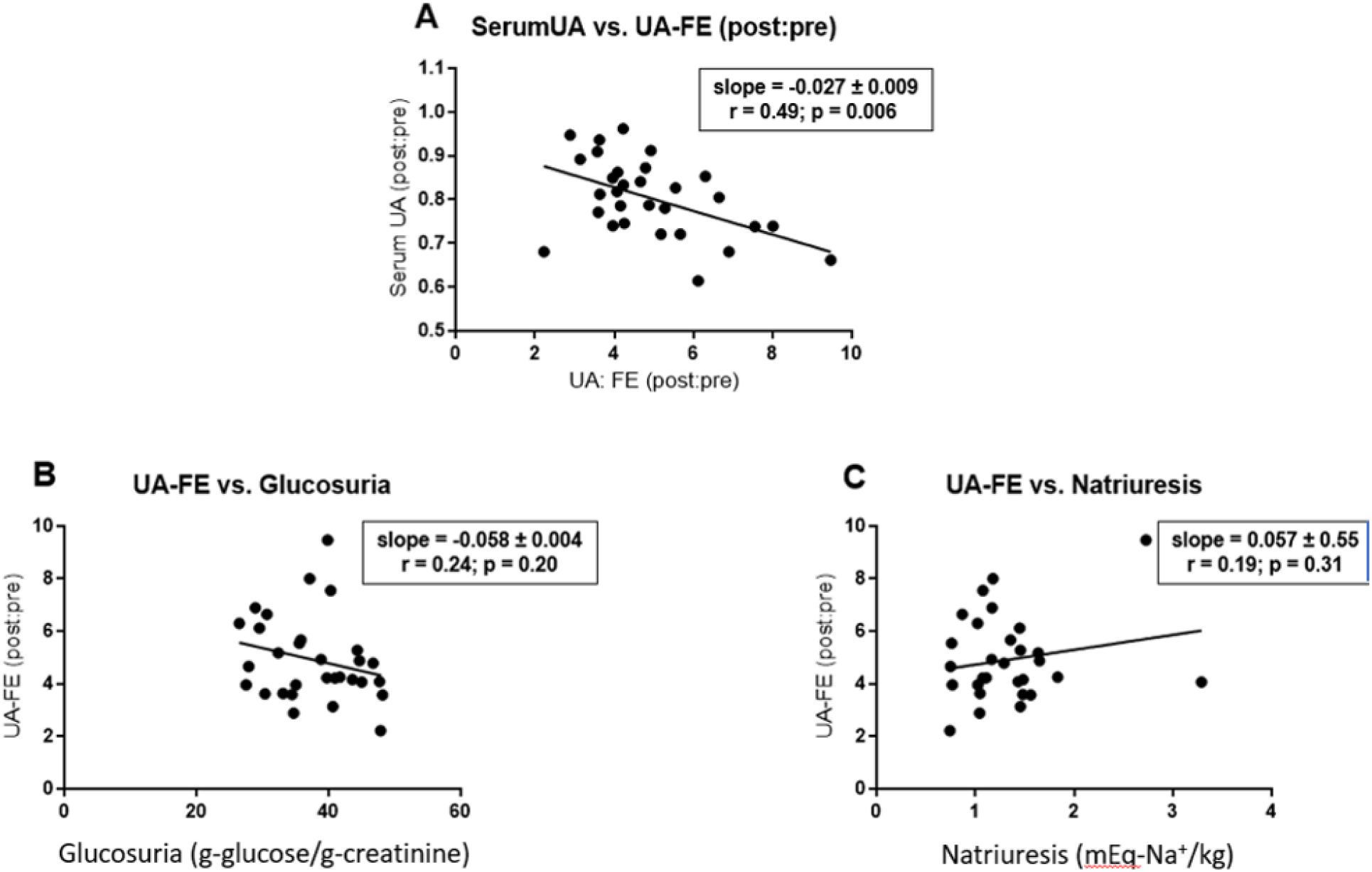
Canagliflozin-induced changes in serum uric acid and fractional excretion of uric acid. Panel A represents a plot of the effect of canagliflozin on serum uric acid level as a function of the canagliflozin-induced increase in fractional excretion of uric acid. Both parameters are presented as ratios of post-canagliflozin levels divided by pre-canagliflozin levels. Panels B and C represents a plot of canagliflozin-induced fractional change in fractional excretion of uric acid as a function of canagliflozin-induced glucosuria (grams of glucose per gram of creatinine; panel B) or canagliflozin-induced natriuresis (expressed as a ratio of post-canagliflozin values divided by pre-canagliflozin values; panel C). We estimated slopes for the least-square lines and correlation coefficients using data analysis programs provided in Excel. p-values were calculated using GraphPad Prism software.

### 3.7 Pilot pharmacogenomic investigation of genetic variants in three candidate genes (SLC2A9, SLC5A4, and SLC5A9)

This pilot study was originally designed to investigate possible association of three candidate genetic variants with pharmacodynamic responses to an SGLT2 inhibitor: nonsense variants in *SLC5A4* (rs62239058; p.E139X) and *SLC5A9* (rs850763; p.E593X) and a missense variant in *SLC2A9* (rs1689079; p.V253I). *SLC5A4* and *SLC5A9* encode two homologs of SGLT2 (SGLT3 and SGLT4, respectively) ^29, 30^. *SLC2A9* encodes GLUT9, a transporter reported to exchange glucose for uric acid ^31^.

Because the clinical trial was terminated early when it became apparent that the clinical trial would not meet its recruitment targets within the available time and budget, the study was underpowered to detect association of genetic variants with drug responses. In any case, none of the three genetic variants was associated with a statistically significant difference when compared to the control group. (Table S2). We cannot, however, exclude the possibility that these variants might exhibit significant associations in a substantially larger study with greater statistical power.

## DISCUSSION

Following NIH’s initiative to improve scientific rigor and reproducibility ^13^, we leveraged this pilot study to validate the experimental protocol supporting our ongoing genome-wide association study (GWAS) to identify genetic variants associated with pharmacodynamic responses to SGLT2 inhibitors (NCT02891954). Based on quantitative comparisons of creatinine content in independent 24-hour urine collections, we confirmed that research participants successfully provided highly reproducible 24-hour urine collections. We did, nevertheless, observe a statistically insignificant trend toward a decreased mean creatinine content (−2.6%) in the second 24-hour urine collection (Fig. S3). Although not statistically significant, we suspect that this small difference may be real and reproducible. Canagliflozin-induced natriuresis leads to a modest volume contraction as reflected in the 6.4% increase in serum creatinine levels (Fig. 1B). The modestly decreased creatinine excretion during the second urine collection is consistent with the expected transient decrease in urinary creatinine excretion characterizing the transition between two steady-states (before and after administration of canagliflozin). Thus, even the small, statistically insignificant difference between the two 24-hour urine collections probably reflects the known pharmacological effects of canagliflozin rather than experimental variation.

Furthermore, we confirmed that canagliflozin elicited the expected pharmacodynamic responses and obtained information on the magnitude of inter-individual variation, which has informed power calculations for our ongoing GWAS. As expected, a single dose of canagliflozin triggered several statistically significant responses: fasting plasma glucose, serum uric acid, serum creatinine as well as urinary excretion of sodium, glucose, and uric acid (Figs. 1-4; Table 1).

These data are reassuring with respect to the feasibility of our ongoing GWAS for pharmacodynamic responses to canagliflozin. The NIH established an expectation for NIH-supported research to include investigations of sex as a biological variable in 2016 ^14, 32–34^. While there has been debate about the optimal approach to implement this requirement, there is general agreement that sex-related variables may emerge as relevant within the context of a specific research program ^35^. Although some traits (e.g., karyotype sex) may be dichotomous for most individuals, there are many biological traits (e.g., height) that represent continuous variables with considerable overlap between sexes ^35^. We used participants’ self-reported sex as a proxy for karyotype sex. We applied this binary classification as the basis to conduct an exploratory investigation of whether an individual’s sex was associated with pharmacodynamic responses to canagliflozin. In parallel, we compared three approaches to normalizing our data – i.e., expressing drug-induced glucosuria relative to body surface area, body weight, or urinary excretion of creatinine. When expressed as grams of glucose per gram of creatinine, mean data were very similar in both males and females (Fig. 3B). In contrast, our data revealed substantial sex differences in mean quantities of canagliflozin-induced glucosuria when expressed relative to body surface area (Fig. 3A) or body weight (Fig. 3C). For example, canagliflozin induced an average of 37% more glucosuria in males when expressed on the basis of grams of glucose per kg of body weight (Fig. 3C). These data are consistent with weight loss data reported by the sponsors of dapagliflozin at the FDA’s Advisory Committee meeting on July 19, 2011 ^36^. Specifically, 24 weeks of dapagliflozin therapy induced greater placebo-subtracted weight loss in males than in females (2.76 kg versus 1.22 kg); the FDA’s analysis concluded that the differential effect of sex was statistically significant (p=0.048) – albeit there was considerable overlap between males and females with respect to the magnitude of weight loss ^36^. Also, many factors contribute to determining the magnitude of weight loss in individual patients – including, for example, the magnitude of SGLT2 inhibitor-induced compensatory increase in food intake ^37^. Nevertheless, taken together, these data with dapagliflozin are consistent with the hypothesis that the larger magnitude of SGLT2 inhibitor-induced glucosuria in men (expressed per gram body weight) contributes to a larger magnitude of mean drug-induced weight loss.

This case study exemplifies how routine study of sex as a biological variable does not necessarily conflict with the aims of precision medicine as suggested by DiMarco et al. ^35^, but rather enabled us to create an inclusive database containing data obtained in all participants – regardless of their self-reported sex. Our experience demonstrates that investigations of sex as a biological variable may offer opportunities for investigators to learn whether sex-related variables emerge as being relevant in the context of their particular research programs.

This pilot study confirms the feasibility of conducting a pharmacogenomic study focused on pharmacodynamic effects of canagliflozin in healthy volunteers. Based on assessment of creatinine content of 24-hour urine collections, we conclude that participants in our study provided complete collections of the urine produced during a 24-hour time period.

Reproducibility and completeness of urine collections are critical elements of a scientifically rigorous clinical trial. We observed substantial inter-individual variation of our primary outcome (i.e., canagliflozin-induced glucosuria) – varying over a twofold range from ∼25 to ∼50 grams of glucose per gram of creatinine. By restricting our study to non-diabetic individuals with relatively normal estimated glomerular filtration rates, we limited the contributions of two important potential confounders. Inter-individual variation in fasting plasma glucose and creatinine clearance accounted for ∼1.4% and ∼0.5% of the observed variance in canagliflozin-induced glucosuria. Although our participants spanned a wide range of ages (35-82 years old), the variation in age accounted for only ∼5% of the variance in canagliflozin-induced glucosuria.

Finally, our studies of sex as a biological variable demonstrated that the magnitudes of canagliflozin-induced glucosuria were similar in males and females when expressed on a per gram-creatinine basis. Based on our observations, we conclude that >90% of the observed variance in our primary endpoint is unexplained after accounting for contributions of age, sex, renal function, and fasting plasma glucose. Accordingly, our ongoing pharmacogenomic study (NCT02891954) is well positioned to define the contribution of genetic factors to the relatively large residual variation in the pharmacodynamic effect of canagliflozin.

## Data Availability

Data can be shared with qualified academic researcher under the terms of a data transfer agreement to assure protection of privacy of research participants.

## ACKNOWLEDGEMENTS

We gratefully acknowledge the National Institute of Diabetes and Digestive and Kidney Disease for the following research grants: R21DK105401, R01DK108942, T32DK098107, and P30DK072488. We also gratefully acknowledge contributions of the research participants and the skilled staff at the Amish Research Clinic for their critical roles in making this study possible. We are grateful to Mary Pavlovich, Melanie Daue, and Kathy Ryan for their help with database management. Dr. Laura Yerges-Armstrong provided genotype data enabling us to identify individuals to be invited to participate in this genotype-guided recruitment study.

## AUTHORS’ CONTRIBUTIONS

*<u>Conception of the clinical trial and PI for NIH grant (R21DK105401)</u>*: SIT

*<u>Acquisition and analysis of data</u>*: ALB, H-RC, MEM, EAS, SIT, HBW, ZSY

*<u>Establishment of Old Order Amish genotypedatabase</u>*: BDM, ARS

*<u>Preparation of first draft of manuscript</u>*: SIT

*<u>Revising and approving final version of manuscript</u>*: all authors

*<u>Accountability for all aspects of work</u>*: SIT

## Supplementary Appendix

### 1 Inclusion/exclusion criteria

To be eligible to participate in the clinical trial, individuals were required to be of Amish descent, at least 18 years old, and have BMI between 18-40 kg/m^2^. We established the following exclusion criteria:

- Known allergy to canagliflozin
- History of diabetes, random glucose >200 mg/dL, or HbA1c ≥6.5%
- Taking any of the following medications: diuretics, antihypertensive medications, uric acid lowering medications, or other medications judged to interfere with interpretation of results obtained in the clinical trial.
- Diagnosis of significant chronic diseases affecting cardiovascular, gastrointestinal, pulmonary, or renal systems or other diseases judged to interfere with interpretation of results obtained in the clinical trial.
- Seizure disorder.
- Pregnancy (self-reported) or breast-feeding within the past three months.
- Estimated glomerular filtration rate < 60 mL/min/1.73 m^2^
- Hematocrit <35%
- Liver function tests (ALT or AST) greater than two times the upper limit of normal
- TSH outside the normal reference range for the assay.
- Current symptoms of genitourinary infection or two or more genitourinary infections during the prior 12 months.
- History of osteoporosis-associated bone fracture
- History of unhealed foot ulcer

### 2 Power calculations

We based our recruitment target of 110 individuals on power calculations to provide 80% power (α=0.05) to detect a 25% difference (∼1.0 S.D.) in canagliflozin-induced glucosuria or a 55% (∼0.85 S.D.) difference with respect to the magnitude of the canagliflozin-induced decrease in serum uric acid levels.

- 20 homozygotes for the nonsense mutation in *SLC5A4* (rs62239058; p.E139X)
- 20 homozygotes for the nonsense mutation in *SLC5A9* (rs850763; p.E593X)
- 25 homozygotes for the missense variant in *SLC2A9* (rs1689079; p.V253I)
- 45 “control” individuals who were homozygous for the major alleles of all three genes.

### 3 Conduct of Clinical Trial

After potential participants signed informed consent forms, nurses (a) obtained a detailed medical history; (b) measured height, weight, and blood pressure; and (c) obtained blood samples for screening laboratory tests (hematocrit, fasting plasma glucose, serum creatinine, serum sodium, plasma TSH, and HbA1c). Eligible research subjects participated in two home visits:

- <u>Visit #1</u>. At the first study visit, the nurse explained the process. Women of reproductive age underwent home testing to exclude the possibility that they were pregnant. The research nurse obtained baseline blood samples and provided supplies needed for the study including a single canagliflozin tablet (300 mg) and supplies necessary for collection of two 24-hour urine samples. Research participants were instructed to take the canagliflozin tablet exactly 24 hours after initiating the first 24-hour urine collection. Participants initiated the second 24-hour urine collection immediately after completing the first and immediately after taking the canagliflozin tablet.
- <u>Visit #2</u>. The second home visit took place two days after the first visit and 24 hours after the participant had taken the canagliflozin tablet. The research nurse obtained blood samples to assess pharmacodynamic responses to canagliflozin and collected the two 24-hour urine collections. The nurses also obtained information to confirm whether the participant followed the instructions and to elicit information about possible adverse events.

### 4 Clinical Chemistry

Blood samples were obtained by a research nurse at home visits and collected in test tubes as appropriate for each assay: EDTA anticoagulant (purple top tube) for measurement of hematocrit and HbA1c; heparin anticoagulant (green top tube) for measurement of TSH; gray top tubes containing sodium fluoride and potassium oxalate for measurement of fasting plasma glucose; red top tube for collecting serum samples. After placing gray, purple, and green top tubes on ice, blood samples were transported to the clinical laboratory at the Amish Research Clinic (maximum transport time, 2 hours). After centrifugation (3300 rpm for 10 min), plasma/serum was sent on the same day to Quest Diagnostics for assay. Participants were provided with “pee-splitter” containers, which were used for 24-hour urine collections. A boric acid tablet (1 gram; Sigma Aldrich, PN#B2625) was added to one bottle for collecting urine destined for glucose assays; urine collected in the other bottle was used for measurement of uric acid, sodium, and creatinine. Urine samples were placed on ice or in a refrigerator prior to being picked up by a research nurse for transport to the clinical laboratory at the Amish Research Clinic. Urinary creatinine, sodium, glucose, and uric acid were measured at Labcorp.

### 5 Disposition and adverse events

Forty individuals were enrolled in this clinical trial between the dates of July 13, 2015 – April 13, 2016 (Fig. S1). Ten potential enrollees were excluded for the following reasons (Fig. S1): low hematocrit (1), low TSH levels (2), frequent genital infections (1), pregnancy (1), history of myocardial infarction (1), giant platelets (1), shortness of breath (1), diabetes (1), or antihypertensive medications (1). The remaining 30 participants completed the study. There were two adverse events of mild to moderate severity (Fig. S1). Prior to taking the study medication, one participant experienced an accident while driving a horse and buggy. Another participant experienced loose stools two days after having received a single dose of canagliflozin (300 mg). Both adverse events were judged as unlikely to have been related to participation in the clinical trial (Fig. S1).

**Figure S1.**
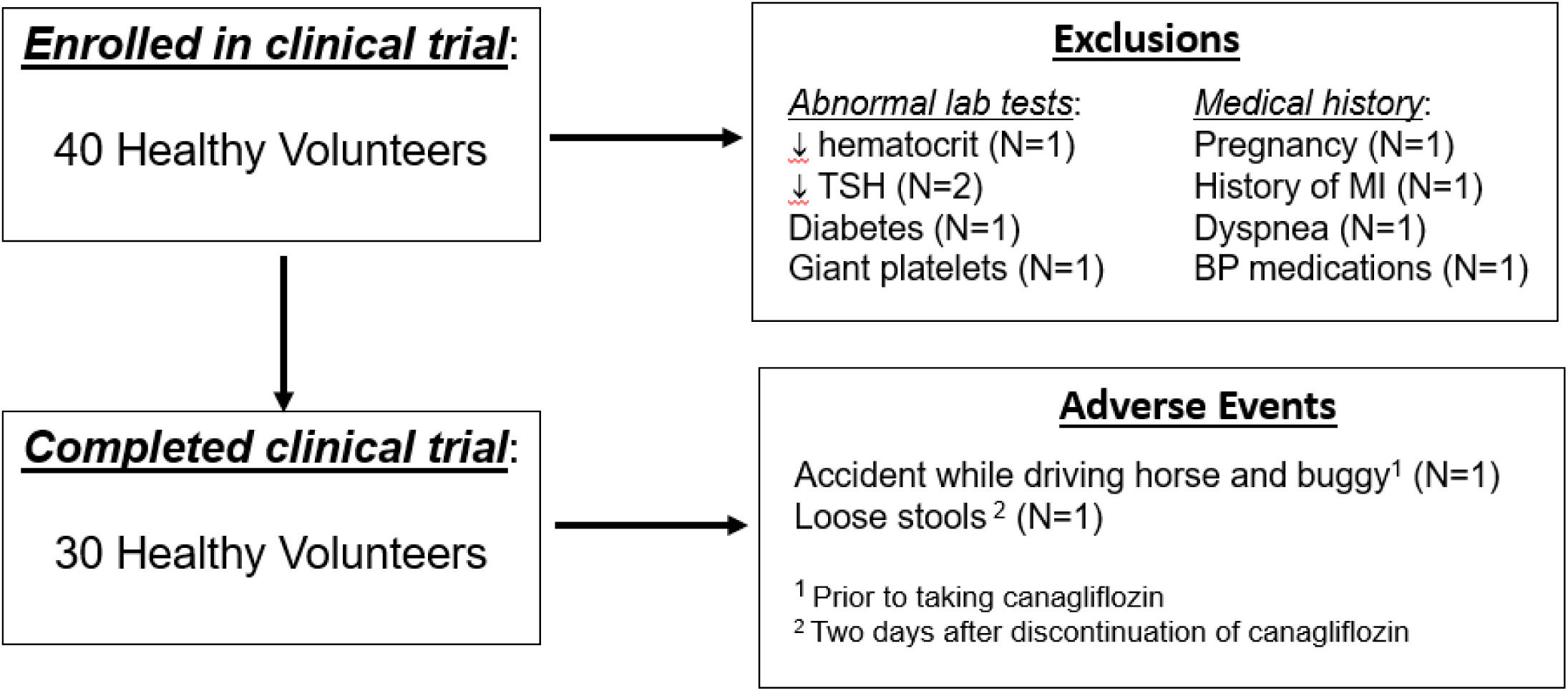
CONSORT diagram: disposition and adverse events.

**Table S1.** Demographics and baseline characteristics of study population.

| **Mean ± SEM (range)** | **Males** | **Females** | **Total** |
| --- | --- | --- | --- |
| **N (sample size)** | 13 | 17 | 30 |
| **Age (years)** | 51.6 ± 3.3 | 62.5 ± 3.4 | 57.8 ± 2.5 |
| **BMI (kg/m<sup>2</sup>)** | 26.5 ± 0.8 | 29.1 ± 1.4 | 28.0 ± 0.9 |
| **Fasting plasma glucose (mg/dL)** | 93.5 ± 2.5 | 92.0 ± 1.8 | 92.6 ± 1.5 |
| **HbA1c (%)** | 5.56 ± 0.08 | 5.81 ± 0.05 | 5.70 ± 0.05 |
| **Serum creatinine (mg/dL)** | 0.80 ± 0.03 | 0.68 ± 0.02 | 0.73 ± 0.02 |
| **Serum uric acid (mg/dL)** | 4.78 ± 0.28 | 4.06 ± 0.18 | 4.38 ± 0.17 |
| **eGFR (mL/min/1.73 m<sup>2</sup>)** | 102 ± 4 | 92 ± 3 | 96 ± 3 |
| **Hematocrit (%)** | 44.4 ± 0.6 | 40.2 ± 0.7 | 42.0 ± 0.6 |

### 7 Pilot pharmacogenomic investigation of genetic variants in three candidate genes (*SLC2A9*, *SLC5A4*, and *SLC5A9*)

This pilot study was designed to investigate possible association of three candidate genetic variants with pharmacodynamic responses to an SGLT2 inhibitor: nonsense variants in *SLC5A4* (rs62239058; p.E139X) and *SLC5A9* (rs850763; p.E593X) and a missense variant in *SLC2A9* (rs1689079; p.V253I). *SLC5A4* and *SLC5A9* encode two homologs of SGLT2 (SGLT3 and SGLT4, respectively) ^1, 2^. *SLC2A9* encodes GLUT9, a transporter reported to exchange glucose for uric acid ^3^.

Renal tubular epithelial cells contain a sufficient number of glucose transporters to accomplish near-complete reabsorption of all the glucose molecules filtered in the glomerulus under normal physiological conditions. SGLT2 is located in the most proximal S1 segment of the proximal tubule and mediates reabsorption of ∼90% of the filtered glucose load under normal physiological conditions ^4^. SGLT1 is located downstream in the S3 segment of the proximal tubule and mediates reabsorption of the ∼10% of the filtered glucose load that escapes SGLT2- mediated reabsorption under normal physiological conditions. In SGLT2 inhibitor-treated patients, SGLT1 has sufficient capacity to compensate partially for the impact of SGLT2 blockade. As a result, the usual doses of SGLT2 inhibitors induce urinary excretion of only 30-50% of the filtered glucose load ^4, 5^, albeit there is substantial variation among individual patients with respect to the magnitude of urinary glucose excretion in response to SGLT2 inhibitors.

In designing this clinical trial, we considered the possibility that two other renal tubular transporters (in addition to SGLT1) might also contribute to reabsorbing glucose when SGLT2 is inhibited by a drug: SGLT4 and GLUT9. These two transporters were selected as candidates in our pilot study because of the existence of relatively common, functionally significant genetic variants in the genes encoding the two transporters: a nonsense mutation in *SLC5A9* (encoding SGLT4) and a missense variant in *SLC2A9* (encoding GLUT9) ^6, 7^. SGLT4 is a homolog of SGLT2 (encoded by *SLC5A2*) that cotransports sodium along with several hexoses including fructose, mannose, glucose, or 1,5-anhydroglucitol ^2^. Among other locations, SGLT4 is reported to be expressed in the S1 and S2 segments of the proximal tubule ^8^. We hypothesized that the loss-of-function mutation in *SLC5A9* (rs850763; p.E593X; minor allele frequency = 0.15) might promote canagliflozin-induced glucosuria by diminishing SGLT4-mediated glucose reabsorption. GLUT9 (encoded by *SLC2A9*) is a member of the SLC2-family of glucose transporters that also includes GLUT1, GLUT2, and GLUT4 among others. GLUT9 is reported to mediate exchange of glucose for uric acid ^3^. A missense variant (rs1689079; p.V253I; minor allele frequency = 0.16) has been identified, which is associated with increased urinary excretion of uric acid and decreased levels of uric acid in serum ^6, 7^. Some publications have proposed that GLUT9 may mediate the uricosuric effect of SGLT2 inhibitors with the increased concentration of glucose in renal tubular fluid driving counter-transport of uric acid ^4, 9, 10^. We hypothesized that the p.V253I variant might be associated with an alteration in canagliflozin-induced uricosuria and/or canagliflozin-induced glucosuria.

*SLC5A4* encodes SGLT3, a homolog of SGLT2 that has lost the ability to transport glucose but has retained the ability to transport sodium ^11–13^. SGLT3 is expressed in the small intestine, where it has been hypothesized to function as a glucose sensor by virtue of its activity as a glucose-regulated sodium transporter ^14, 15^. Because SGLT3 has also been reported to be expressed in kidney ^15, 16^, we hypothesized that a loss of function mutation in *SLC5A4* might enhance canagliflozin-induced natriuresis and/or diminish canagliflozin-induced glucosuria by diminishing SGLT3-mediated sodium reabsorption. We did not observe any trends suggesting that homozygosity for this nonsense mutation in *SLC5A4* (rs62239058; p. E139X; minor allele frequency = 0.08) is associated with alterations in canagliflozin-induced glucosuria or uricosuria. We did, however, observe a statistically insignificant trend (p=0.07) toward an increase in canagliflozin-induced natriuresis – consistent with the presumed role of SGLT3 as contributing to mechanisms of renal tubular reabsorption of Na^+^ ^1, 13, 16^.

The clinical trial was terminated early when it became apparent that we would not meet our recruitment targets within the available time and budget. Nevertheless, we analyzed data to explore whether there might be preliminary trends suggesting possible association of pharmacodynamic responses with any of the three genetic variants. Our primary outcome was glucosuria – a pharmacodynamic biomarker for glycemic efficacy and weight loss. The four genotype groups exhibited mean rates of urinary glucose excretion in the range from 36.9-39.1 g-glucose per g-creatinine (Table 3). None of the three genetic variants was associated with a statistically significant difference when compared to the control group.

We confirmed previously published observations ^6, 7^ that the p.V253I variant in SLC2A9 (rs1689079) was associated with decreased mean levels of serum uric acid levels (3.7 ± 0.27 versus 4.6 ± 0.20 mg/dL in the present study; p=0.02). A previous analysis in the Old Order Amish ^7^ estimated an effect size of 0.44 ± 0.06 mg/dL per variant allele, which is similar to the magnitude of the difference observed in the present study (0.9 mg/dL) for homozygotes carrying two variant alleles. Thus, our current observations appear to have reproduced previously published data. We did not, however, detect statistically significant association of any of the genotypes with alterations in fractional excretion of uric acid – either at baseline or in response to canagliflozin (Table S2). It is likely that our clinical trial lacked the statistical power to detect genetic associations unless effect sizes were quite large.

**Table S2.**
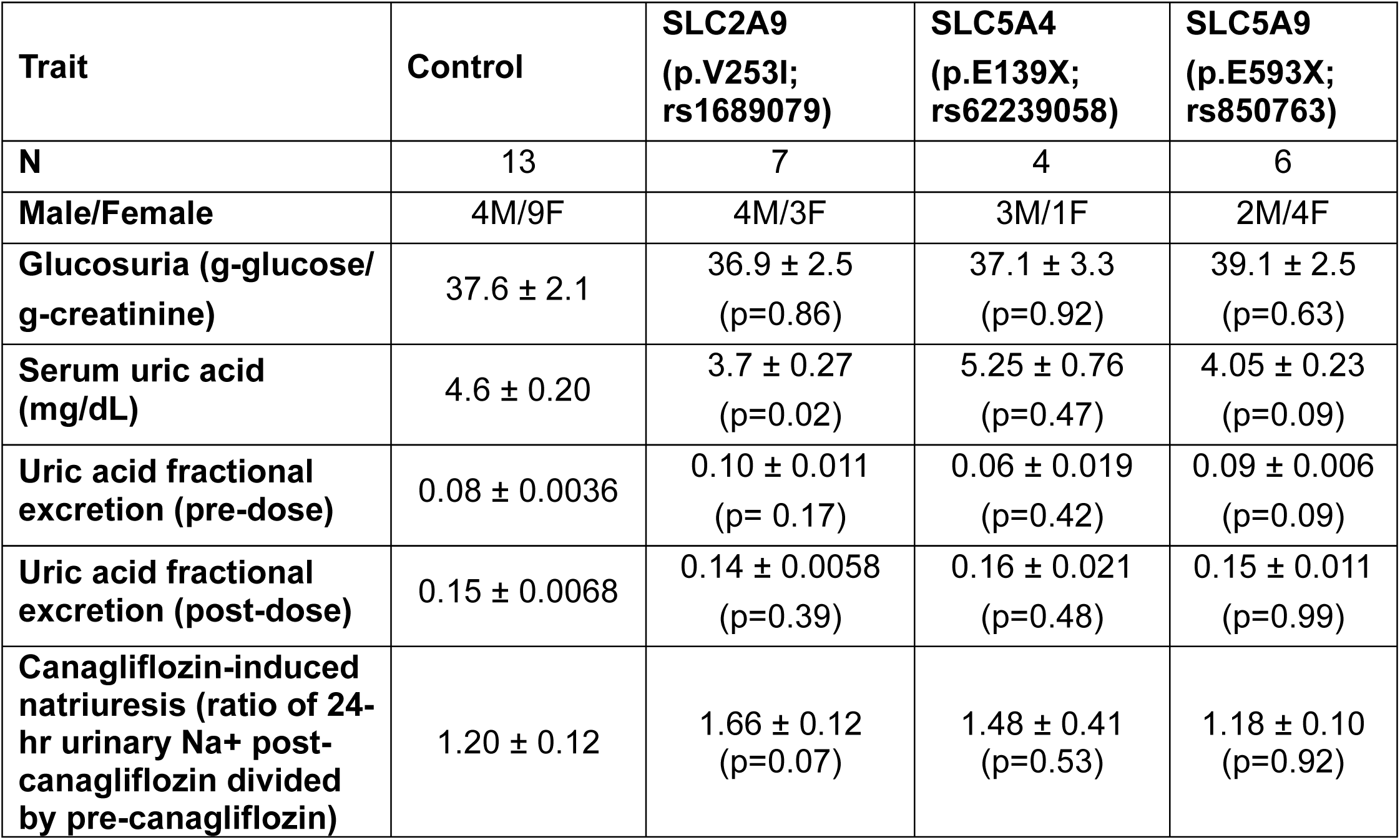
Selected data stratified according to genotype. Data are presented as means ± SEM. p-values were calculated relative to the control group using t-tests for unpaired data without correction of multiple comparisons.

| Trait | Control | SLC2A9<br>(p.V253I;<br>rs1689079) | SLC5A4<br>(p.E139X;<br>rs62239058) | SLC5A9<br>(p.E593X;<br>rs850763) |
| --- | --- | --- | --- | --- |
| **N** | 13 | 7 | 4 | 6 |
| **Male/Female** | 4M/9F | 4M/3F | 3M/1F | 2M/4F |
| **Glucosuria (g-glucose/**<br>**g-creatinine)** | 37.6 ± 2.1 | 36.9 ± 2.5<br>(p=0.86) | 37.1 ± 3.3<br>(p=0.92) | 39.1 ± 2.5<br>(p=0.63) |
| **Serum uric acid**<br>**(mg/dL)** | 4.6 ± 0.20 | 3.7 ± 0.27<br>(p=0.02) | 5.25 ± 0.76<br>(p=0.47) | 4.05 ± 0.23<br>(p=0.09) |
| **Uric acid fractional**<br>**excretion (pre-dose)** | 0.08 ± 0.0036 | 0.10 ± 0.011<br>(p= 0.17) | 0.06 ± 0.019<br>(p=0.42) | 0.09 ± 0.006<br>(p=0.09) |
| **Uric acid fractional**<br>**excretion (post-dose)** | 0.15 ± 0.0068 | 0.14 ± 0.0058<br>(p=0.39) | 0.16 ± 0.021<br>(p=0.48) | 0.15 ± 0.011<br>(p=0.99) |
| **Canagliflozin-induced**<br>**natriuresis (ratio of 24-**<br>**hr urinary Na<sup>+</sup> post-**<br>**canagliflozin divided**<br>**by pre-canagliflozin)** | 1.20 ± 0.12 | 1.66 ± 0.12<br>(p=0.07) | 1.48 ± 0.41<br>(p=0.53) | 1.18 ± 0.10<br>(p=0.92) |

**Figure S2.**
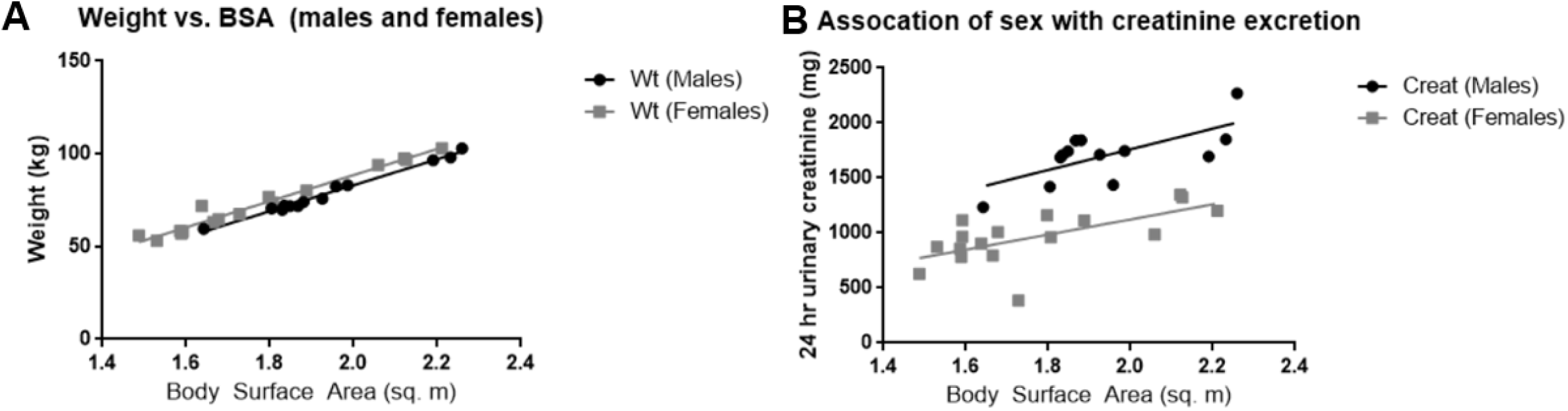
Correlation of urinary Na+ excretion before and after administration of canagliflozin. *<u>Panel A</u>*. Individual participants’ body weights are plotted as a function of calculated body surface areas (r=0.99 for both males and females). *<u>Panel B</u>*. 24-hour urinary creatinine excretion is plotted as a function of body surface area (r=0.68 for males and r=0.65 for females). Data for males are represented by black circles and for females with gray squares.

**Figure S3.**
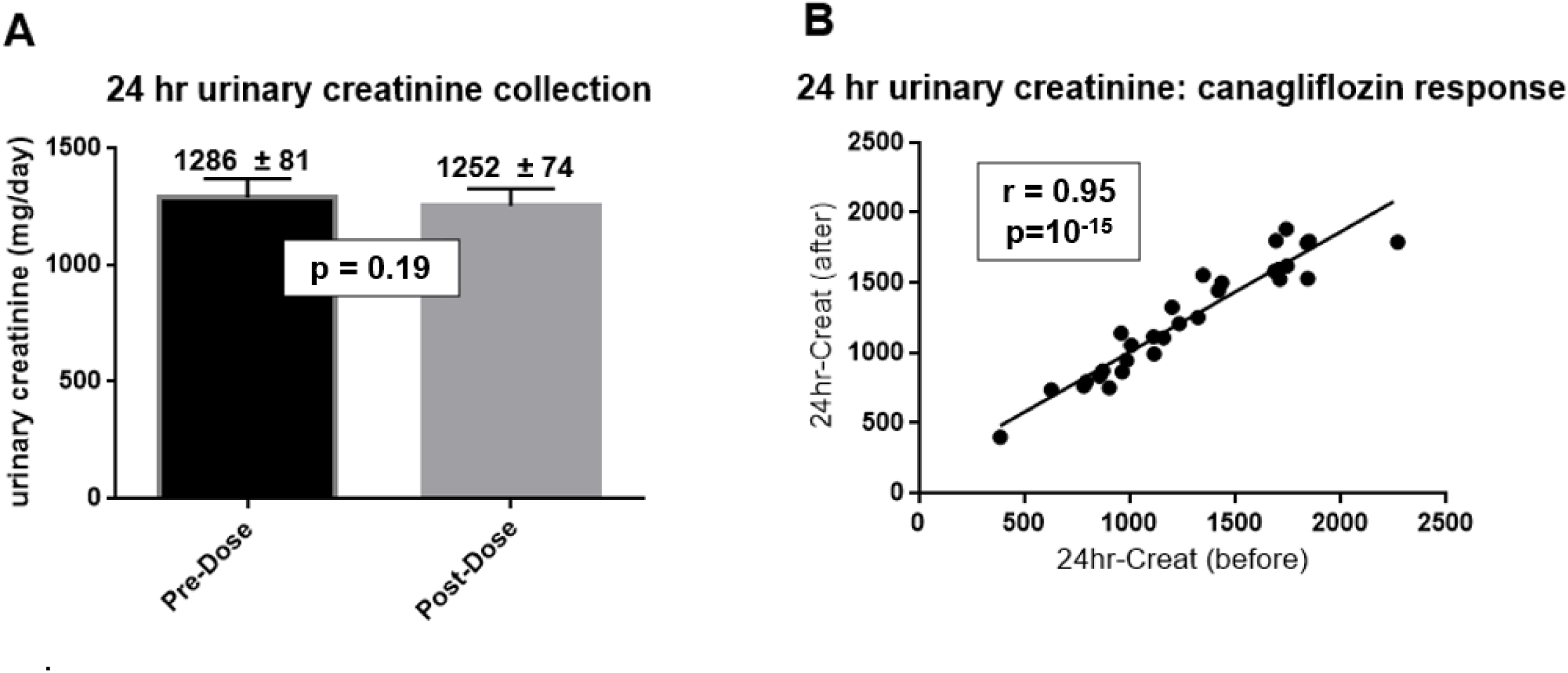
Reproducibility of 24-hour urine collections. After participants collected the first 24-hour urine collection, they received one tablet of canagliflozin (300 mg, p.o.). Immediately thereafter participants collected a second 24-hour urine collection. Data are presented as mean ± SEM (N=30)

**Figure S4.**
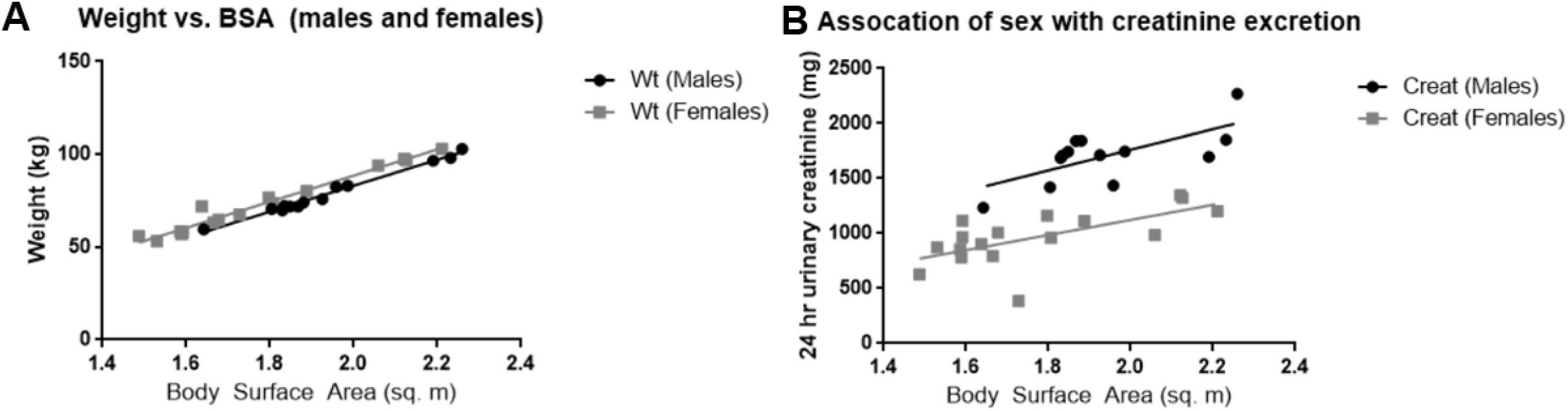
Correlations among indices of body size (body weight, body surface area, and daily creatinine production): impact of sex as a biological variable. *<u>Panel A</u>*: Individual participants’ body weights (kg) are plotted as a function of body surface area (sq. meters) with males indicated by blue circles and females by red square symbols. Data are presented as mean ± SEM (N=30). *<u>Panel B</u>*: Individual participants’ 24-hour urinary creatinine excretion values are plotted as a function of body surface area (sq. meters) with males indicated by black circles and females by gray square symbols. Linear regression yielded equations of **Y**=939**X**-118 (males) and **Y**=686**X**-250 (females) where **Y** represents 24-hour urinary creatinine (mg/day) and **X** represents body surface area (sq. meters). For a body surface area of 1.8 m^2^, the equations predict 24-hour urinary creatinine excretion rates of 1572 mg/day (males) and 985 mg/day (females) – i.e., ∼60% higher for males.

**Figure S5.**
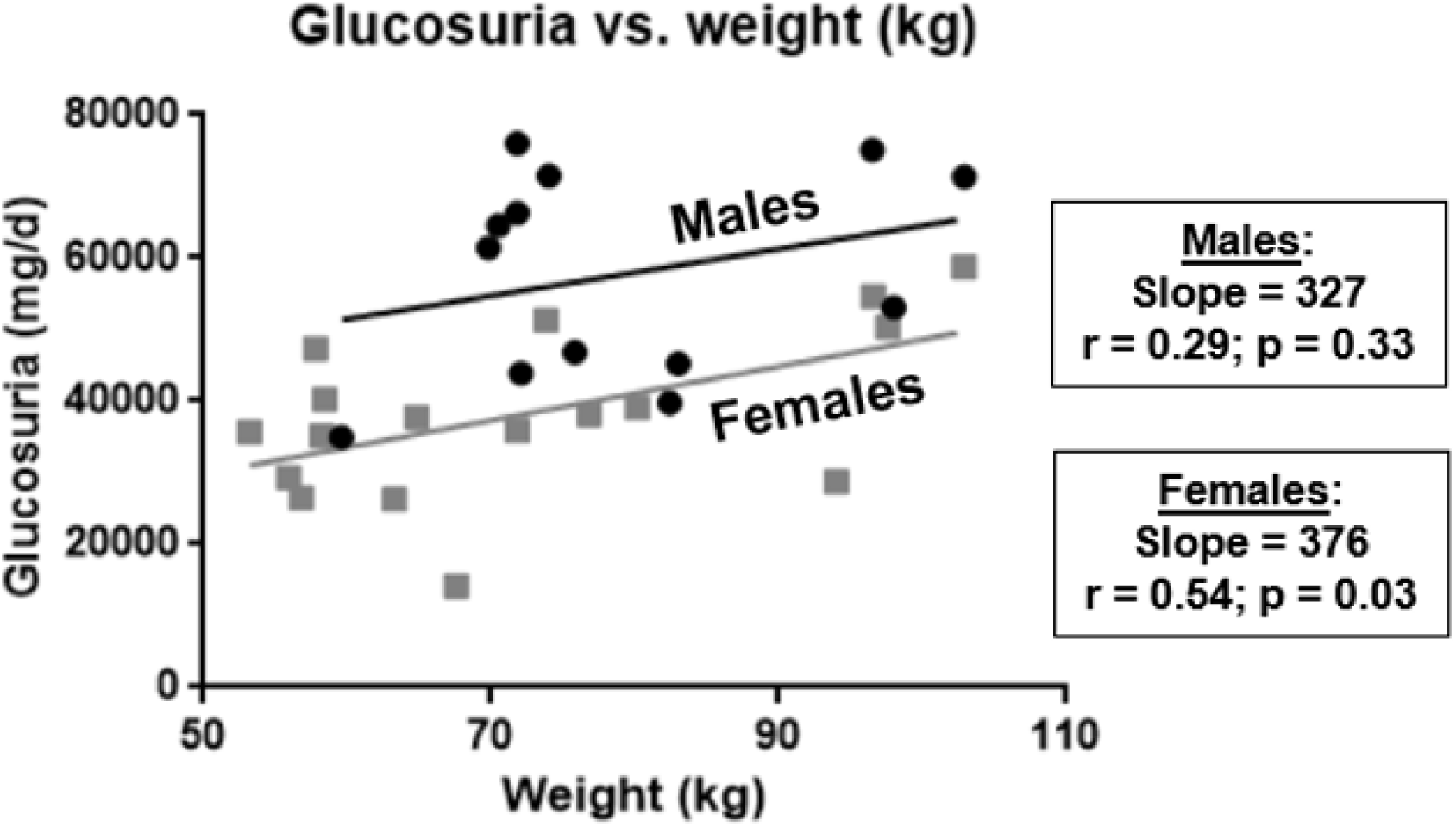
Sex differences with respect to the magnitude of canagliflozin-induced glucosuria when normalized relative to body weight. Canagliflozin-induced glucosuria (mg/kg/d) was plotted separately for males (black circles) and females (gray squares) as a function of body weight (kg).

**Figure S6.**
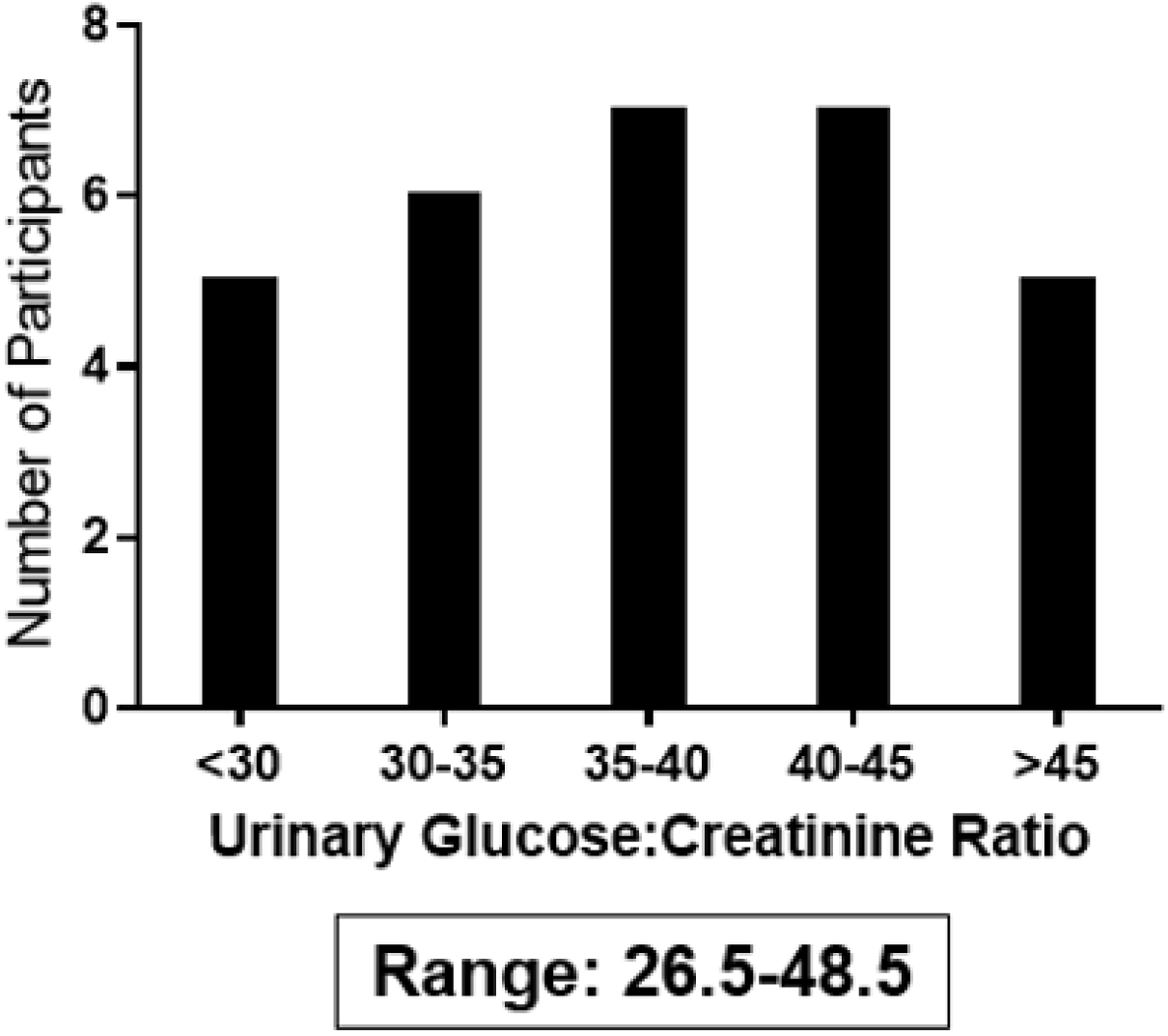
Inter-individual variation in 24-hour urinary glucose excretion. Using data from Figs. 5 and 6, we constructed a histogram depicting the number of individuals (y-axis) with various levels of 24-hour urinary glucose excretion (grams of glucose per grams of creatinine). The observed data ranged between 26.5 to 48.5 g-glucose/g-creatinine.

**Figure S7.**
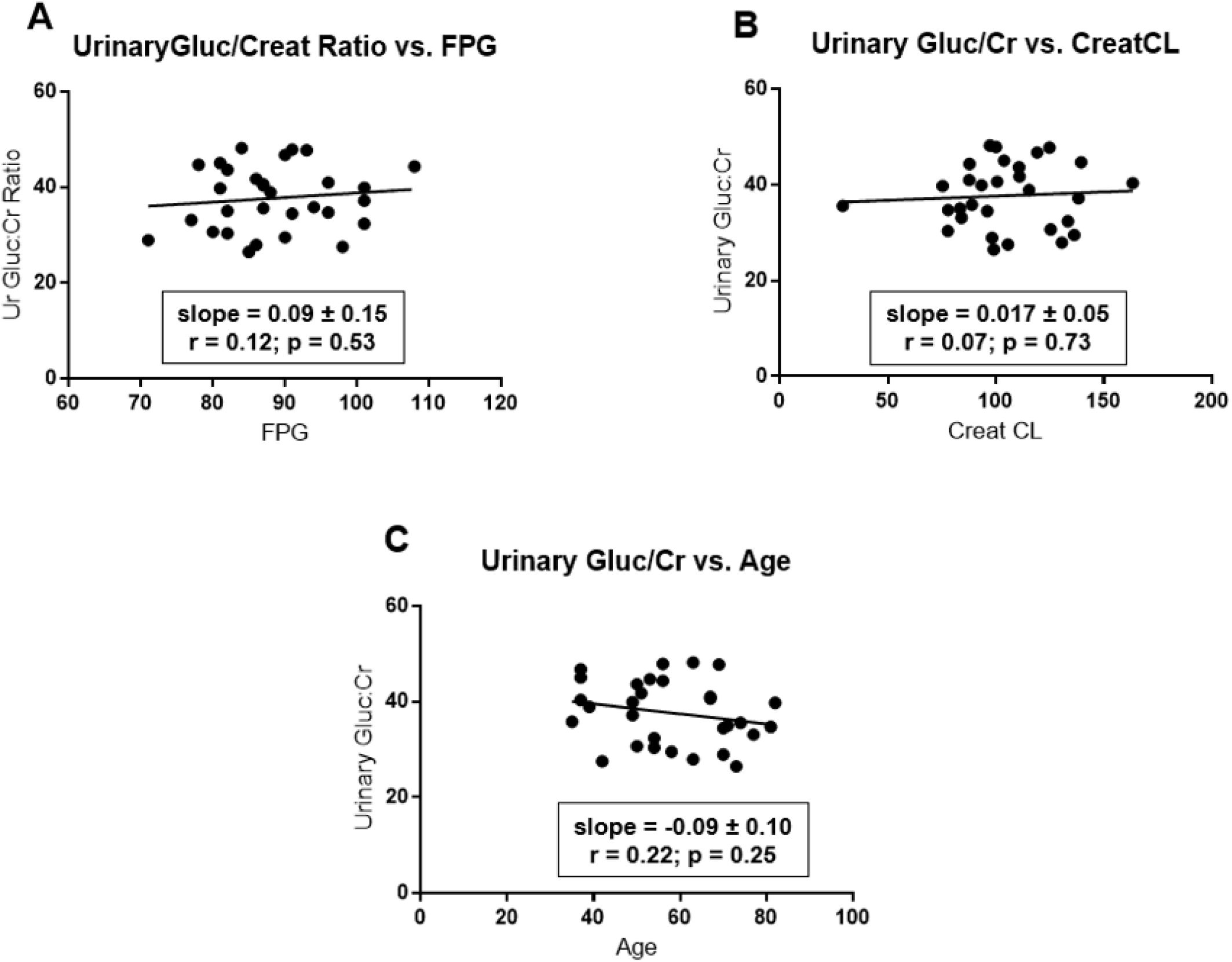
Correlation of urinary glucose:creatinine ratios in 24-hour urine collections with selected parameters. Data from Figs. 5 and 6 were analyzed to assess correlation of 24-hour urinary glucose excretion rates with fasting plasma glucose (panel A), creatinine clearance rates (panel B), and age (panel C). We estimated slopes for the least-square lines and correlation coefficients using data analysis programs provided in Excel. P-values were calculated using GraphPad Prism software.

**Figure S8.**
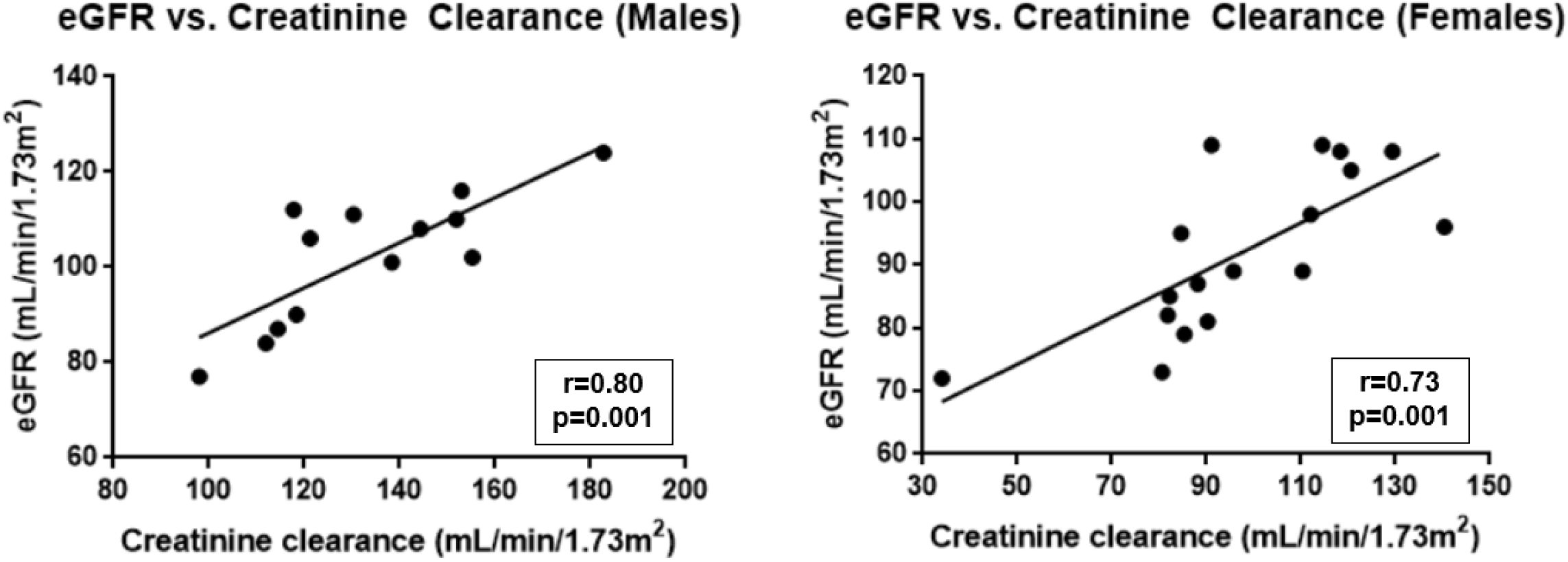
Correlation of estimated GFR (eGFR) with measured creatinine clearance. eGFR (calculated by Quest Diagnostics) is plotted on the y-axis as a function of measured creatinine clearance. We calculated creatinine clearance based on lab data obtained prior to administration of canagliflozin: serum creatinine and 24-hour urinary creatinine excretion. *<u>Left panel</u>*, males; *<u>right panel</u>*, females.

